# Primary health care improvement in Benin: Cocreating an evidence-informed policy framework to guide the practices of primary care physicians

**DOI:** 10.1101/2024.03.11.24304111

**Authors:** Kéfilath Bello, Bart Criel, Jean-Paul Dossou, Djimon Marcel Zannou, Jan De Lepeleire

## Abstract

**Introduction:** In Benin, policies for guiding the practices of primary care physicians (PCPs) are sparse and incomplete. This leads to sub-optimal use of these relatively rare human resources and reduces their potential contribution to people’s health and well-being. In this study, a policy framework guiding PCPs’ practice in Benin is developed in cooperation with key stakeholders.

**Methods:** The cocreation was a long-term process spread over several years, culminating in a two-day workshop in Cotonou (Benin’s capital city) in October 2022. The core principles of a successful cocreation process were respected: a clear definition of goals, the participation of all relevant stakeholders, including community members, and evidence-informed discussions.

**Results:** The resulting policy framework includes four overarching dimensions: first, the objectives assigned to PCPs in Benin; second, their roles and related activities; third, their professional identity; and fourth, the governance arrangements guiding their practices. The policy framework provided concrete recommendations for these dimensions based on the Benin context and international evidence.

**Conclusion:** The cocreation process was instrumental in developing an evidence-informed and consensual policy framework guiding PCPs’ practices in Benin. The framework may be relevant for other West African countries but must be customised to each country’s context.

## Introduction

In Benin, the burden of disease is still high, with, e.g. a maternal mortality ratio in 2020 of 523 deaths per 100,000 live births versus 531 and 223 deaths per 100,000 live births in the whole of Africa and in the world, respectively [1]. Similarly, in 2021, the country had a high under-five mortality rate at 83.52 deaths per 1,000 live births, and its health services coverage index was among the lowest in Africa, at 38 out of 100 in 2021 [1].

This situation can be considerably improved with the effective implementation of Primary Health Care (PHC) [2], which includes integrated and quality health services to meet people’s health needs (known as “primary care”), addressing the broader determinants of health through multisectoral policy and action, and empowering individuals, families, and communities to take charge of their health [3]. An effective PHC system requires a sufficient, well-distributed and well-performing health workforce [4,5], especially at the interface between the community and the health services system – i.e. first-line health services - where most of the population health needs should be addressed [3,4].

For decades, the provision of primary care in Benin, as in many African countries, has been delegated to non-physicians because of the limited number of physicians [6,7]. However, the ratio of physicians in Benin has gradually increased, from 0.6 to 1.5 physicians per 10.000 inhabitants between 2010 and 2017 [8,9]. This increase, combined with the development of the private sector and limited access to postgraduate training opportunities, increased the presence of physicians working at the primary care level. Because of their more elaborate university training than non-physicians, one would naturally assume that the increased presence of Primary Care Physicians (PCPs) would improve primary care delivery. Indeed, some studies, including one in Benin, indicated that PCPs could potentially improve primary care performance in Africa [10–12]. However, a scoping review [13] conducted by our team in 2020 on PCPs’ practices in Sub-Saharan Africa showed that this improvement should not be taken for granted. When poorly regulated, PCPs may actually fail to improve care quality. They also struggle to ensure certain key principles of PHC, such as accessibility, continuity of care, patient-centeredness, or responsiveness to people’s health needs [13]. Moreover, in contexts where non-physicians already substantially contribute to primary care delivery [6,7], PCPs need to find their place in the ecosystem of primary care in order to avoid tensions, redundancies or even deviances, for instance, engaging in activities situated outside their competencies in order to prove their “superiority” [14,15]. Another contextual particularity is that PCPs in Benin need to work with fewer resources than their peers in other parts of the world. In 2020, Benin’s current health expenditure per capita was 32 US dollars. In comparison, Belgium, Cuba, Brazil, South Africa, and Thailand spent 5009, 1186, 701, 490, and 305 US dollars in the same year [1].

All these issues call for well-thought policies steering PCPs’ practices to optimise their contribution to PHC. Indeed, one of the four strategic levers recommended by WHO’s operational framework for PHC is to develop sound policy frameworks that will support actions in vital areas [16]. Policy frameworks are documents that describe a set of goals, procedures, and principles that might guide more detailed policies and interventions on a given issue [17]. In Benin, a sound policy framework is timely, given that PCPs are still a relatively rare and expensive resource, even if they are on the rise. A *laissez-faire* policy would thus lead to inefficiencies in utilising this resource, as suggested by a study conducted in 2020 in four health districts in Benin [18]. However, Benin lacks, like many other African countries, a clear, explicit and contextualised policy framework to support PCPs [13].

The present study aims to close this gap by developing, in collaboration with key stakeholders, a policy framework to guide the PCPs’ practices in Benin. PCP’s practices are defined by the services provided by the PCPs, their individual characteristics, the features of their workplace, and the team (if any) within which they operate. The research question that guided us was: “what should be the content of a policy framework guiding the practices of PCPs in Benin, taking into account the specificities of the country’s context?”.

## Methods

### Definition and principles of cocreation

To ensure that this policy framework is well connected to Benin’s health policy and systems processes, we used a cocreation approach for its development. Cocreation is a process where relevant stakeholders (researchers, policymakers, practitioners, end-users) work together to generate evidence on a given topic, diagnose related problems and their root causes, and formulate potential solutions [19,20]. As such, cocreation helps ensure that the evidence produced is relevant to local priorities and that the solutions proposed and the decisions taken are contextualised, connected to ongoing policy processes, feasible, acceptable, and endorsed by many stakeholders.

In the literature, cocreation is related to (and sometimes used interchangeably with) several concepts. For instance, in its series on increasing the impact of health research through coproduction of knowledge, the BMJ used the term “coproduction” to designate a collaborative model of research where researchers work in partnership with knowledge users to identify a problem and produce knowledge, sharing power and responsibilities from the start to the end of the research [21]. Another related concept is “policy dialogue”, which is an iterative process considering both the technical and political aspects of a problem, involving evidence-based and politically sensitive discussions, including a broad range of stakeholders, and having a concrete purpose or outcome in mind [22]. Thus, policy dialogue and cocreation share many similarities, especially because they both seek to ensure that the best available evidence informs policy and practice decisions and imply a sustainable partnership between researchers, policymakers, health systems users and other stakeholders [21–24].

No matter the terminology used, a cocreation process should follow some principles to be successful. First, the goals of the cocreation should be clearly defined from the onset and understood by all stakeholders [20,22,25]. The cocreation process should also be inclusive, with trusted relationships and shared power among the stakeholders [22,24,26,27]. The discussions and decisions during cocreation should be informed by evidence [22,28,29], with the stakeholders’ experiences and perspectives as valued as the evidence provided by scientific research [24,28,30].

The discussions should also be well-prepared, ensuring that the relevant stakeholders are present and that the evidence needed is shared timely with the stakeholders in adequate formats [28,31]. Finally, a successful cocreation needs skilled and dedicated facilitators and relevant tools (relevant collaborative tools, knowledge products, etc.) [28,30,31].

### Conceptual framework and steps of the cocreation process

Based on the principles and steps described above, we developed a conceptual framework to guide our cocreation process (see Figure 1).

**Fig 1.**
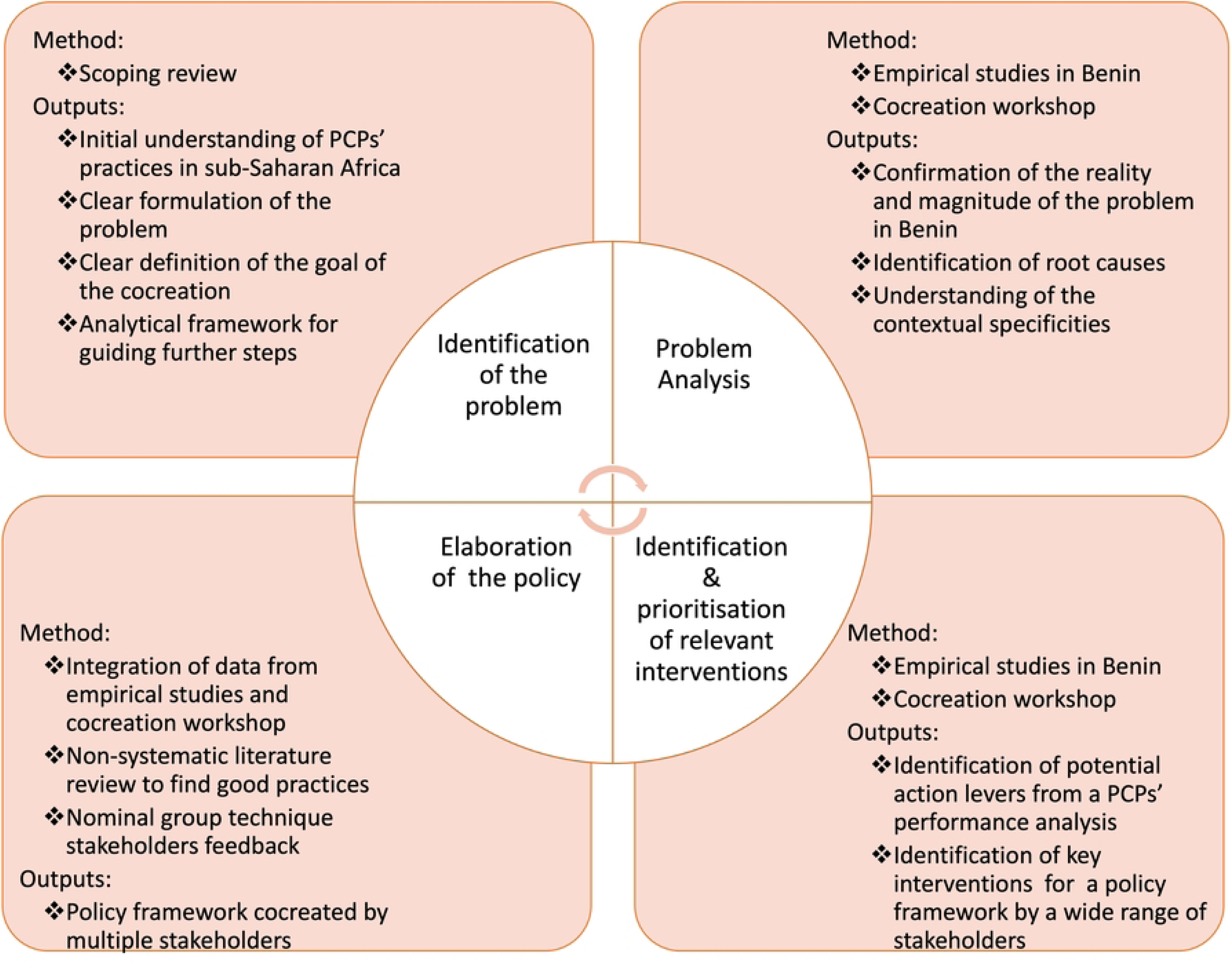
Conceptual framework for the cocreation process.

At the step of problem identification, we conducted a scoping review [13], which helped us better understand the practices of PCPs in sub-Saharan Africa. We were then able to properly formulate the problem we were trying to solve, i.e. “the PCPs in most sub-Saharan African countries are not reaching their potential for improving PHC and there is little policy guidance for guiding their practices”. We could also clearly formulate the goal of the cocreation, which is to develop a policy framework to guide the PCPs’ practices for PHC improvement in Benin. Another output of the scoping review was an analytical framework for analysing the PCPs’ practices in further stages and guiding the development of the policy framework.

For the problem analysis step, we conducted three mixed methods studies between December 2019 and July 2022 in five purposively selected health districts in Benin: Cotonou 2 and 3 (Cot 2-3), Cotonou 5 (Cot 5), Ouidah-Kpomassè-Tori (OKT), Parakou-N’Dali (PN) and Nikki-Kalalé-Pèrèrè (NKP). The first study described and categorised the PCPs’ practices in Benin [18]. The second study learnt from the PCPs’ experiences with the COVID-19 pandemic to further understand the strengths and weaknesses of their practices, especially during crises [10]. In the third study, we analysed the performance of 8 interesting cases of PCPs’ practices, selected based on their performance levels and other characteristics, to understand the factors that influence these practices [32]. These three studies allowed us to confirm the reality and magnitude of the problem in Benin (i.e., the sub- optimal performance of the PCPs), identify the root causes, and understand the contextual specificities of the problem. The problem analysis was finalised during a cocreation workshop described below.

At the step of identification and prioritisation of relevant interventions, we used information from the third empirical study described above and information from the cocreation workshop (see S1 file). The empirical study on the PCPs’ performance helped us to find potential levers of actions that could be considered for the policy framework. The cocreation workshop was a two-day workshop in Cotonou organised by the research team. In this workshop, a wide range of stakeholders from Benin (see table 1) worked collaboratively to analyse the issues identified in the PCPs’ practices in Benin, propose solutions, and reflect on various aspects of the policy framework. Two participants from Togo and Guinea were invited, and the discussions extended to the West African context. However, most recommendations focused on Benin’s context.

**Table 1:** Overview of the stakeholders engaged during the cocreation process.

| Cocreation activities<br>(Period of the activity) | No. of participants | Profile of participants |
| --- | --- | --- |
| Study 1 (5 December 2019 to 15 August 2020) | 155 | • PCPs who participated in a survey, of whom 21 also participated in IDI |
| Study 2 (18 April 2020 to 15 August 2020). | 90 | • PCPs who participated in a survey, of whom 14 also participated in IDI |
| Study 3 (19 April 2022 to 7 June 2022) | 157 | • PCPs (IDI participants): n=9<br>• Non-physician PHC workers (IDI participants): n=9<br>• Non-physician PHC workers (FGD participants): n=54<br>• Patients (IDI participants): n= 16<br>• Other community members (FGD participants): n=63<br>• Local health authorities (IDI participants): n=6 |
| Cocreation workshop (25 October 2022 to 26 October 2022) | 26 | • Policymakers from the Ministry of Health (n=5, including 1 from Togo)<br>• Policymakers from the Ministry of Social Affairs (n=1)<br>• Civil society (n=3)<br>• Development partners (n=4)<br>• Health providers organisations/all cadres from the private sector (n=3)<br>• Health providers organisations/non-Physicians (n=1)<br>• Health providers organisations/physicians (n=3, including 1 from Guinea)<br>• Local health authority (n=4)<br>• Medical schools (n=2) |
| Informal stakeholders-engagement activities | N/A | • Policymakers at the national level (n=6, including 5 from the Ministry of Health and 1 from an international organisation)<br>• Study participants cited above |

The last step was the elaboration of the policy framework. After the workshop, the research team produced a draft of the policy framework, integrating the workshop results and those of the empirical studies (S2 file). The team also used a non-systematic literature review (including several policy documents, intervention reports and peer-reviewed articles) to look for good practices to enrich the policy framework. The draft policy framework was then discussed among a group of researchers and public health practitioners using a nominal group technique [33]. The refined framework obtained after this exercise was shared with all participants at the workshop and policymakers for feedback.

### Ensuring the quality of the cocreation process

We strived to follow the principles of a successful cocreation process.

The scoping review was helpful to ensure a good definition of the problem and the goal of the cocreation. The research team elaborated a concept note that clearly explained the workshop’s objectives and was shared with the stakeholders before the workshop.

To ensure inclusivity, trusted relationships, and shared power, we set up a long-term stakeholder engagement from the outset of the cocreation process. This includes several in-person discussions on research methodologies and preliminary results with PHC policymakers at the national level and sharing the results and relevant resources with the PCPs participating in the various studies. The results were shared in the format of short oral presentations, sometimes supported by a PowerPoint, an evidence brief with infographics, and research syntheses sent via email and WhatsApp to various stakeholders. This stakeholder engagement helped us gather regular feedback and build ownership of the process. An important aspect to mention here is the fact that the third empirical study provided an opportunity to gather the viewpoints of the patients and community members. These viewpoints were shared with the participants of the cocreation workshop. Table 1 provides an overview of the stakeholders engaged throughout the cocreation process.

The cocreation process was also informed by evidence as the discussions among the stakeholders and the development of the policy framework was largely informed by the empirical studies conducted in Benin, an extensive literature review, and experiences shared by the workshop participants.

Finally, a team of facilitators, led by KB and supervised by BC, JDL and MDZ, carefully prepared the cocreation workshop. During the workshop, we combined oral presentations, plenary discussions, and group work. Various techniques were used to gather participants’ ideas and prioritise policy options, including brainstorming, facilitated group discussions, and a world café (where groups rotated among three discussion tables akin to café settings to examine various issues). The key issues discussed are presented in Table 2. We also allocated more than half of the workshop time to cocreation activities and discussions.

**Table 2:** Key issues discussed during the cocreation workshop.

| Themes of the group discussions | Guiding questions/instructions | Facilitation technique used |
| --- | --- | --- |
| Root causes analysis of the problematic issues identified in the PCPs' practices in Benin | Based on the studies' results presented in the plenary, identify the root causes of the issues identified concerning the PCPs' practices in Benin—use of the "5 whys" approach. | Group discussions facilitated by research team members. The facilitators started by asking why the identified issue was observed and then, from the answers of the participants, drilled down into successive "whys" to the most distal cause. |
| Objectives, values and principles of the PCPs' practices in West Africa | Based on the root causes analysis and your knowledge of the West African context, answer the following questions:<br>What should be the objectives of the PCPs' practices in West Africa?<br>Which outcomes should we expect from a first-line health facility with a PCP?<br>What should be the performance criteria of the PCPs in West Africa?<br>Which values should guide the PCPs' practices in West Africa?<br>Which principles should West African PCPs follow? | Group discussions facilitated by research team members |
| Roles, activities, governance, and professional identity of the PCPs in West Africa | Questions series 1:<br>What should be the composition of a primary care team at first-line health facilities? | World café, with three tables dealing each with a questions series. Three groups of participants were formed, and each group was assigned to a table to |
|  | • Which services should a first-line facility with PCPs offer?<br>• What are the roles of a PCP within a primary care team?<br>• What are the roles of the other members of the primary care team?<br>Questions series 2:<br>• Does a physician need postgraduate training to work as a PCP?<br>• How do we manage the careers of PCPs to retain them in the primary care system?<br>• How do we ensure good accountability of PCPs toward the community and other key stakeholders?<br>Questions series 3:<br>• How do we ensure good regulation of the PCPs' practices in West Africa to achieve the expected outcomes?<br>• What are the minimum resources required for helping PCPs achieve the expected outcomes? | reflect on the corresponding questions.<br>After 20 minutes, the groups changed tables and worked on the subject of the new table. After two rotations, each group could discuss all the questions. A facilitator and a rapporteur were assigned to each table. The rapporteur took note and presented a summary during the plenary. |

### Ethical considerations

We obtained ethical approvals for all original research, which results were used for the cocreation process. Specifically, the cocreation workshop was conducted under the ethical approval N° 0513/CLERB-UP/P/SP/R/SA issued by the local ethics committee for biomedical research of the University of Parakou and the ethical approval N° 1545/21 issued by the Institutional Review Board of the Institute of Tropical Medicine of Antwerp. We obtained a written informed consent from the participants to the empirical studies. Participants in the workshop all freely contributed to elaborating the policy framework and signed an attendance list. We managed all the data with strict confidentiality. We avoided putting the names of the participants (other than the research team members and the officials who provided public remarks) in all shared documents.

## Results

### Overview of the framework

The overall organisation of the policy framework is modelled on the classic organisation of several health systems frameworks. Indeed, the starting point of the policy framework is the framework produced from the scoping review which is inspired by:

- the Donabedian model [34] which provided the general architecture (structure, process, results)
- various health systems analysis frameworks, including the WHO building blocks of the health system [35] and the health systems dynamics framework of Van Olmen et al. [36]: since PCPs operate within a system, these frameworks guided us for a comprehensive and systematic identification of the items that could influence the performance of PCPs.

This initial framework was iteratively refined and enriched throughout the cocreation process. The resulting policy framework (Figure 2) includes the following overarching dimensions: first, the objectives assigned to the PCPs in Benin; second, the roles and activities of the PCPs; third, the professional identity of the PCPs in Benin; and fourth, the governance elements of the PCPs’ practices. We described each dimension and subdimension of the policy framework in the following sections. We enrich these descriptions with verbatims/quotes illustrating the meaning participants give to some dimensions. The comprehensive viewpoints of the workshop participants are documented in the workshop report (S1 file).

**Fig 2.**
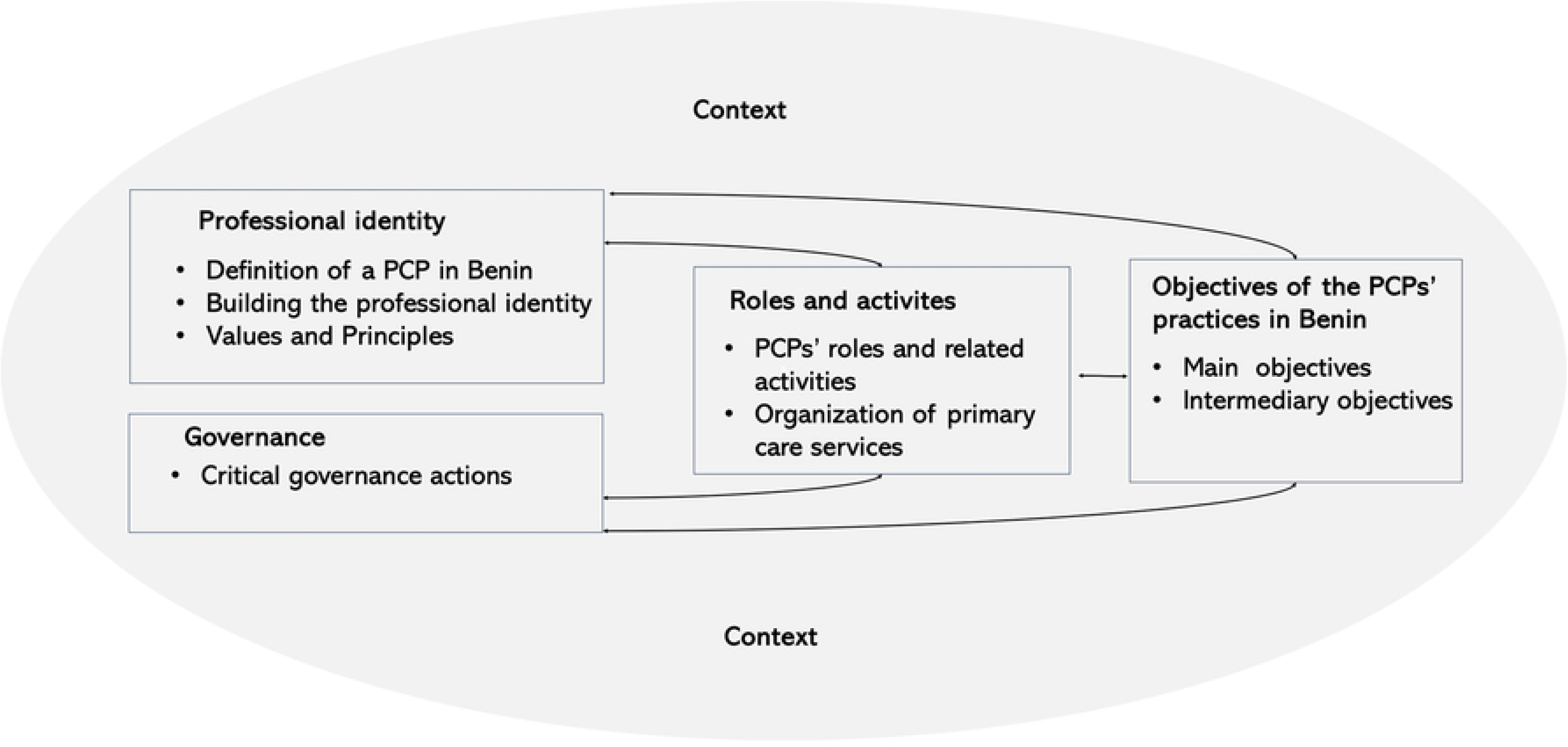
Overview of the policy framework.

**Fig 3.**
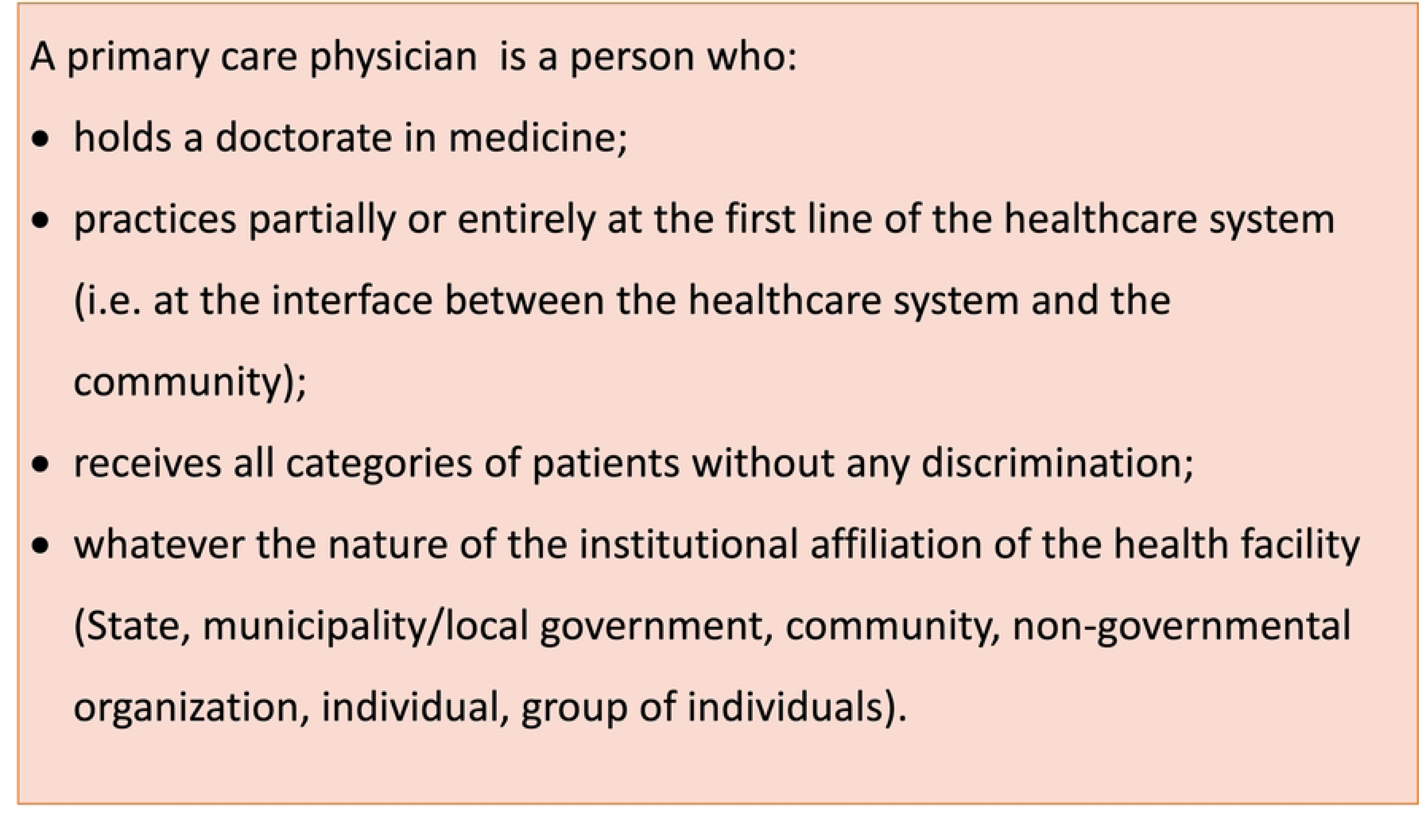
Definition of a primary care physician in Benin.

### Objectives assigned to the PCPs in Benin

#### Main objectives

Workshop participants agreed that the overall goal of the PCPs’ practices in Benin is “ to improve people’s health, well-being, and socio-economic conditions, including reducing morbidity and mortality”. However, achieving this goal depends on more than just the PCPs’ practices. Therefore, stakeholders retained four specific objectives for the PCPs’ practices in Benin.

The first is to ensure the quality of care offered at the primary care level (first-line and community). The following dimensions of quality of care were agreed upon:

- Patient-centeredness: Most participants in the studies and cocreation workshop stated that the care provided by the PCPs should meet the patients’ and the community’s needs and preferences. They have particularly insisted on a good welcome of the patients and good listening, enabling patients to express themselves and PCPs to understand the patient’s concerns and build a trusting relationship with them. Furthermore, stakeholders stressed the need to consider the psychological aspects when dealing with a patient.
- Effectiveness means that the PCPs are able to make the correct diagnosis and that the care provided follows sound standards. Stakeholders also insisted on the fact that effective care should provide relief to patients regarding their health issues;
- Safety, meaning that the care provided should not harm the people receiving it;
- Integration, meaning that the PCPs and their team are able to provide the necessary services to the patients and community in a coordinated way, including health promotion, preventive and curative services.
- Continuity, including a long-term follow-up of the patient beyond one-time consultations.

The quotes below exemplify the participants’ perception of this first objective.

> *“The doctor needs to take into account the patient’s psychology, so that the patient feels relieved…. Even if they [the doctors] have their own moods, they should leave all that behind when they are in front of the patients to welcome us, put us at ease and then, automatically, we are half cured” (community member, FGD, Southern Benin, 2022)*.

> *“The doctor needs to be a good listener, ask more in-depth questions to make a good diagnosis, and take concrete actions to resolve the problem of the diagnosis they have made. Then, thank the patient, give them an appointment so that they can come back, and I think that if they do that, hey, 95% of customers will say that this doctor is good” (primary care worker, FGD, Northern Benin, 2022)*.

The second objective is to ensure the accessibility of healthcare to the population served. This includes:

- Geographical accessibility, which can require community outreach activities;
- Cultural acceptability, whereby the PCPs and their team try to adapt to the culture of the population served and avoid inappropriate attitudes and practices;
- Financial accessibility: the population complained about the high cost of PCPs’ services, and the workshop participants recommended that the PCPs strive for financial accessibility through cost control or seeking financial protection solutions with community members;
- Availability of the PCPs and their team when the community needs them;
- Timely access;
- Equitable access, meaning that all people should have access to the care provided by the PCPs and their team, especially those who need it most.

The third objective is to ensure a sound referral system for the patients. This entails:

- A timely and safe referral of the patients who need it to the hospital,
- Good communication and coordination with other levels of care (community, various hospitals) and providers outside the primary care team (specialists, psychologists, social workers, etc.) in the local health system.

> *“We have to ensure that they (the patients) are referred when the technical conditions here do not allow to take care of them, that they are referred on time, and that the mortality rate is almost zero.”* (PCP, in-depth interview, Northern Benin, 2022).

The fourth objective is to ensure the satisfaction of patients and the community. Based on the stakeholders’ statements, this satisfaction means that:

- The services provided by the PCPs should solve the problem that brought the patients to them.
- The patients and the community should be at ease with their healthcare providers.

> *“In terms of the doctor’s performance, the patient’s satisfaction comes first. And when the patient is satisfied, and you give them an appointment, they will come back. It may be chronic illnesses that require follow-up; you will always see them, and they will come. This facilitates collaboration, and at the same time, the health centre is better attended (local health authority, in-depth interview, Northern Benin, 2022)*.

#### Intermediary objectives

Three intermediary objectives were also retained.

The first of these intermediary objectives is to contribute to the availability and sound performance of the primary care team. For this objective, PCPs are expected to ensure that the primary care team members are effectively present when the patients need them (thus no absenteeism) and have proper behaviour. PCPs are also expected to ensure that the primary care team members (including themselves) get continuous medical education and are able to provide good quality care effectively. To achieve this objective, the PCPs can also advocate with authorities and other key stakeholders to acquire an adequate primary care team and the necessary resources to strengthen this team’s capacities.

The second intermediary objective is to support the management of health facility resources. This includes supporting the mobilisation and management of financial resources, the availability of adequate infrastructure and equipment, effective medicine supplies and sound hygiene in the facility. However, there is no full consensus on this particular objective. While local health authorities and PCPs from the public sector support it, other stakeholders, especially non-physicians and the PCPs operating on a private basis, emphasised that time invested in management tasks may go at the expense of patient care. Moreover, the performance analysis results indicate that delegation of the facility management to a professional supports PCPs’ performance and secures time for patient care.

The quotes below illustrate both the first and the second intermediary objectives.

> *“Apart from quality, people need to be able to see whether their presence is organising the work or the service… Are the staff acting correctly? Because if they are there and there is a mess, that means they are not doing their job. If they (the doctor) are there, the organisation must be good, we must have staff who are better than the others. And at the same time, if the number of patients increases, does the revenue increase? If they are there and the patients increase, and on the other hand they don’t make sure that the revenue is well managed, that’s another problem. So, in most of these centres where there is a doctor, illicit sales (of medicines) have practically stopped”.* (local health authority, in-depth interview, Northern Benin, 2022).

> *“Because if she (the doctor) doesn’t work well, will we go to the center again and see that everything is clear? It’s like you have your own room, and every morning when you wake up, you don’t sweep it; everything will be a mess. But when everything is clean and well organised, you know that the person in charge is doing their job properly.* (community member, FGD, Northern Benin, 2022).

The third intermediary objective is to contribute to building a health information system supporting patient care and decision-making. PCPs are thus meant to properly collect, analyse, and use the necessary data for decision-making and improvement. They are also expected to encourage other primary care team members to collect, analyse and use the data where appropriate.

### Roles and activities

#### PCPs’ roles and related activities

Six roles were defined for Beninese PCPs: care provider, guarantor of the quality of care, interface between population and other actors within the health system, planner and coordinator of health activities targeting a given population, health educator and community advocate, and leader of the care team.

As a **care provider**, the PCP’s key activities include the provision of medical consultations to the patients. However, almost all stakeholders agreed that, because of their limited number, the PCPs in Benin may focus on complex cases (especially non-communicable diseases) and emergencies. As a result of the latter, many stakeholders suggested extending the package of services provided by a first-line health facility with a PCP. However, others were wary of such an extension because it may lead to higher costs and overlaps with the hospital activities. It was agreed that further reflections are needed to decide which services should be added when a PCP is in a first-line health facility. Stakeholders also strongly recommended that PCPs integrate prevention and health promotion into the care they provide.

The **guarantor of quality of care role** is one of the roles in which the stakeholders find the most added value in the PCPs, compared with the other primary care workers. Therefore, PCPs are expected to oversee and enhance the quality of the care provided to the patients and community. They may thus train and supervise other primary care team members. They should also lead or participate in relevant quality assessment and quality improvement activities, for instance, clinical audits, regular discussions with their team about the patients they received, etc.

As **the interface between populations and other actors**, the PCP is responsible for screening the population and referring them to other professionals as early as possible when needed. They are the anchor point of the patient in the health system. They organise the referral of the patients when needed, and they coordinate the actors and interventions needed by the patients and the community.

As **planner and coordinator of health activities for a given population**, the PCP is responsible for planning the health activities with the primary care team according to the needs and the national health policy. They are also responsible for coordinating the implementation of these activities. This role will require the PCPs to perform non-clinical activities such as budgeting, reporting, etc.

As **health educator and community advocate**, the PCP should actively seek good relationships with the community, including community leaders. This can be done through dialogues with community members. PCPs are also expected to provide relevant information to maintain the good health of community members through, for instance, sensitisation campaigns or other health communication approaches. The PCPs should also listen to the community, identify its needs and advocate for fulfilling them.

As **the leader of the care team**, the PCP takes responsibility for achieving the objectives assigned to the care team. This includes seeking resources and guiding other team members to achieve shared goals. While the PCPs are not the only ones who can be leaders in a primary care team, the empirical data showed that PCPs who were given a leadership mandate and the necessary autonomy to apply it enhanced the teams’ performance. They succeed even more when they adopt a person-centred leadership style, valuing and respecting other team members and promoting shared objectives, values and collaboration within the team.

The following quotes highlight some of these roles.

> *“I was saying that the role of a doctor is to see the patient from the very beginning of the illness, to ensure that the pathology doesn’t develop into something serious, and that the patient is treated as soon as possible… The PCPs play a vanguard role in public health. At the first line, when we see cases that seem epidemic to us, we raise the alarm and inform the authorities”* (PCP, in-depth interview, Northern Benin, 2022).

> *“They (the PCPs) have to mobilise their teams, dynamise them and make sure that they don’t go off the rails… They have to encourage work well done, while giving greater responsibility to the players in the health system.”* (local health authority, in-depth interview, Northern Benin, 2022)

> *“If there is a new disease coming, for example, she can raise awareness among the population so that they change their behavior to avoid it… You know that the doctor is there to coordinate, so when something like this happens, she informs her team and sends her team out into the field to raise awareness.”* (Community member, in-depth interview, Northern Benin, 2022)

#### Organisation of primary care services

Three themes emerged related to the organisation of primary care services.

The first one is that PCPs in Benin should not work alone or in silos. They should instead work in teams with other primary care team members. This teamwork implies task-sharing, cooperation and mutual learning. Through task-sharing, each cadre of the primary care team would take on the tasks where they could have added value. According to the data gathered, the added values of the PCPs in Benin include:

- Complex diagnosis and procedures (such as care for chronic diseases and complications)
- Training and coaching health workers with less knowledge and skills
- Relationships and negotiations with community and health leaders and with specialist physicians.

As for non-physicians (especially nurses-practitioners), the stakeholders (both in the empirical studies and the workshop) see their added value for:

`o` Regular follow-ups of patients (they appear to be more available and less intimidating to the patients)
`o` Long-term relationships with patients
`o` Task related to their specific training (e.g. nursing, social support, etc.).

However, there can be flexibility in the organisation of the team, with, for instance, the possibility of non-physicians providing medical consultations for simple cases when needed and if they have the necessary skills. All team members should cooperate around the needs of the patients and learn from each other, as stated by a community member below:

> *” The second thing I want to say, whether it is the assistant, the nurse or even the doctor, what we want to tell you and tell all the people of Benin is that they are all health workers, and it is not suitable for them to fight over a patient. It is important that there is a mutual understanding because the nurse has, let’s say, their own brain, the assistant has theirs, and so does the doctor. So, it’s important that when a patient comes in, and the illness is a bit serious, everyone consensually agrees on a diagnosis to decide what needs to be done to the patient to regain their health.* (Community member, FGD, Northern Benin, 2022).

The second theme related to the services organisation is community engagement. The PCPs and other primary care team members should seek partnerships with the community leaders and key stakeholders within the community. This can be achieved through regular communication, collaboration and participation in key community development activities.

Finally, an effective and up-to-date referral system should support the primary care team. Stakeholders recommended the establishment of a mechanism or a platform for smooth referrals and enabling contact with specialists. This referral platform should include all needed services and providers (even beyond the health sector). The platform is physical (network of providers) and digital (online platform). There were also recommendations for innovation and improvement of the current referral system (for instance, adapting the system to consider the spontaneous networks currently used by PCPs, the urban context, etc.).

### Professional identity

In the policy framework, we defined professional identity as “the set of attributes, values, principles, motives, and experiences that PCPs and other stakeholders use to define PCPs in their professional work”. This definition was adapted from the one of Caza et al. [37]. The policy framework includes three sub-dimensions of professional identity: the attributes of a PCP in Benin, the values and principles that should guide PCPs’ practices, and recommendations for strengthening PCPs’ professional identity.

## Definition of a PCP in Benin

The PCPs’ definition (see Figure 2) was adopted by the workshop’s participants (S1 file) and validated during subsequent discussions with the stakeholders.

## Values and principles

The values highlighted in the framework are the following: respect for people, empathy, altruism, community orientation, humility, professionalism, and integrity.

Respect for people entails that PCPs should listen to their patients and their team members. They should also respect them and respect the patient’s rights and cultural identity.

Through empathy, the PCPs will ensure that patients are well treated and get the best experience throughout their care pathway.

Altruism implies that the PCPs will pursue the well-being of the patients and community first instead of personal gains.

By embracing humility as a value, the PCPs are expected to be open-minded, adaptable, creative, and proactive. They should also show a spirit of teamwork and cooperation.

Professionalism implies that the PCPs will seek competence and work with dedication and quality. They will also demonstrate leadership and display good communication skills.

Community orientation demands that the PCPs assess and respond to the needs of the community served.

With the integrity value, the PCPs will avoid malpractices (such as ransomware) and respect the best standards of care.

According to the stakeholders that contributed to the creation of this policy framework, these values and principles should guide the PCPs’ practices and the practices of the primary care team.

> *“Respondent: The first thing is integrity; a doctor has to be honest, a doctor has to have integrity..*.

> *Interviewer: What do you mean by being honest?*

> *Respondent: Honesty… Honesty and integrity… It is a value that requires human beings to do things as they should, without trying to round things up for their own well-being or personal needs. For me, a doctor must be honest and have integrity, so it’s the same thing. He needs to give the best and fairest possible response to the patient. A doctor doesn’t lie. A doctor doesn’t juggle; a doctor doesn’t fake things. Also, a doctor has to be empathetic; that is very important. He must learn to be compassionate about society’s problems…”* (PCP, in-depth interview, Northern Benin, 2022).

## Building the PCPs’ professional identity

The stakeholders strongly emphasised that developing and strengthening a specific professional identity of PCPs is essential. This includes the promotion, among the PCPs, other primary care team members and future primary care workers, of the values and principles stated above and explaining and promoting the performance objectives expected from the PCPs.

Another related recommendation is that policymakers and training institutions give PCPs official status. This includes developing postgraduate training, changes in salary, etc. This would facilitate their integration into national policies. It would also increase their motivation and retain them within the primary care practice.

### Governance

Governance is the function of a health system that ensures policy guidance, regulation and coordination of different functions and actors, optimal allocation of resources, and accountability towards the population and all stakeholders [13,36]. In this policy framework, the governance dimension includes ten actions that can support PCPs in their roles and activities and facilitate the achievement of the objectives assigned to them.

## Designing and implementing adequate regulations and policies

For this element, the policy framework recommends that the Ministry of Health, the medical council and other institutions responsible for developing the PCPs’ practices clearly define the rules for PCPs’ training, posting and career development, their roles, their salaries and other financial incentives, etc. This policy framework was cited as a key step for this element. Its recommendations could help to integrate PCPs’ practices better in national health policies and health sector regulatory documents.

## Ensuring an adequate preparation of PCPs

The performance analysis of the PCPs’ practices showed that good preparation of the PCPs prior to or at an early stage of their job appointment is critical. Such preparation will give the PCPs the necessary values, knowledge, attitudes, and skills for practising at the primary care level (and, if needed, in underserved areas). This preparation can be done through undergraduate training, postgraduate training or early field coaching.

## Providing relevant continuing education of PCPs in both public and private sectors

The data largely highlighted the lack of a well-established and systematic continuing education system for the PCPs. This policy framework thus proposes the development of a training curriculum for the PCPs. Training topics suggested by the stakeholders include (but are not limited to) community health issues, health promotion, public health, care for the most frequent medical conditions and emergencies, care of complex health issues, leadership and management, values for the PCPs’ practices, etc.

Another recommendation for this element is establishing a platform for training and scientific exchange among PCPs, other primary care workers, specialists, and other relevant stakeholders. This platform can also enable the PCPs to be in contact with peers and specialists.

## Establishment of a supportive supervision and coaching system

Even though physicians are among the most highly trained cadres of health workers, the empirical data and the stakeholders’ perceptions agreed on the necessity to support the PCPs in their daily work. This support can be provided to PCPs through coaching by experienced and trusted professionals, who will provide continuous guidance for improving performance and dealing with unknown or complex situations. PCPs can also benefit from supportive supervision, including on-the- job visits to monitor and provide feedback on their performance. Coaching and supportive supervision must also include psychological support to PCPs to cope with work-related stress (as data collected during the COVID-19 pandemic emphasised). Supportive supervision and coaching can be provided by senior officials within the health system, experienced PCPs, professional organisations, etc. The empirical data showed that peer support is also valuable and can be achieved through regular discussions, experience sharing, and community-building activities.

## Promoting and strengthening PCPs’ organisations

This element was deemed important because, if stronger than they currently are, the PCPs’ organisations can support the PCPs, promote PCPs’ values and facilitate good excellence among primary care physicians. This promotion of PCPs’ organisations can start with the medical council and existing organisations such as the association of the *“médecins généralistes communautaires”*.

## Providing a multidisciplinary primary care team

Having agreed that the PCPs in Benin should work within a team, the stakeholders recommended defining the minimum composition of a multidisciplinary primary care team depending on the context and the utilisation level of the health facility. Policymakers and key stakeholders should support primary care facilities to get the minimum care team. An ideal composition of a multidisciplinary primary care team in Benin was proposed during the workshop and this includes 1 PCP, 1 midwife, 1 nurse-practitioner, 1 pharmacy clerk, 1 cashier, 1 laboratory technician, 1 social worker, 1 physiotherapist, 1 psychologist, 2 to 3 nurse-aids, 1 cleaning staff, 1 security guard, 1 hygiene officer, 1 administrative and finance staff. However, the stakeholders recognised that having the ideal team will not be feasible in many settings, and some cadres may not be needed full-time.

This is why they suggested promoting the mutualisation of some cadres (for instance, administrative and financing staff or psychologists) between facilities if feasible and needed.

## Ensuring adequate equipment, infrastructure, and consumables

The stakeholders also recommended defining the minimum equipment, infrastructures, and consumables needed, depending on the health facility’s context and utilisation level. The presence of a PCP would increase the complexity of the cases treated and the interventions delivered by the primary care team. The equipment and infrastructure should thus be adapted accordingly. However, the stakeholders strongly recommended carefully assessing the added value of any new equipment so as not to increase costs unnecessarily.

Once the minimum equipment, infrastructures, and consumables are defined, the PCPs and their teams should be supported to get them.

## Design and enforcement of sound accountability mechanisms

Setting up sound accountability mechanisms was highlighted in both the empirical data and the workshop as an important element for enhancing PCPs’ performance. Relevant accountability mechanisms for the PCPs in Benin include:

- Clearly defining and sharing the roles and responsibilities of the PCPs (and primary care team members), the expected performance objectives, and how they will be assessed. The current policy framework can provide a basis.
- Defining and regulating the geographic area or the population to be covered by the PCPs’ practices to guarantee good coverage across the entire Benin population.
- Ensuring the PCPs have the necessary capacities, resources, and autonomy to fulfil their role and achieve the objectives.
- Ensuring a regular assessment of PCPs and related incentives and sanctions
- Ensuring the participation of PCPs in the monitoring of the local health systems’ indicators and various health systems activities in both public and public sectors (for feedback and learning)
- Establishing accountability platforms where PCPs can share their experiences and report to the population (e.g. Local Development Associations, Health Centre Management Committees, etc.).

## Careful design of adequate financing modalities of PCP practices

The financing modalities of the PCPs should ensure a decent salary and the financial sustainability of their practices. This is especially vital in the private sector as the empirical data shows that a lack of financial support may lead the PCPs and their facilities to increase costs or over-prescribe health services. Financial support to the PCPs can include facilitating access to loans to establish a practice, carefully chosen financial incentives or even indirect financial support such as health insurance, financial guidance, or access to professional financial management for the health facility. Finally, the financing modalities of the PCPs can support their accountability if geared towards the results expected from the PCPs and integrated with other accountability arrangements.

## Building a health information system that is supportive to PCPs’ performance

The stakeholders strongly suggested providing a well-performing health information system to support the PCPs’ practices. Indeed, they argued that the PCPs and their team will need a sound health information system to ensure the continuity of care, good referrals and learning about their practices. The stakeholders particularly insisted on a better digitalisation of the health information system, especially patient files, through, for instance, patient data recording or accounting management software. They have also insisted on involving the PCPs and other primary care team members in re-designing these tools and the health information system.

### Context

In this policy framework, the context relates to any element outside the boundaries of a given PCP’s practice. It includes members of other practices, other health workers and services, other parts of the local health system, other sectors (e.g. the social welfare sector), community members, general national policies, the socio-economic and political context, etc.

While this policy framework can serve as general guidance for the PCPs’ practices in Benin, its recommendations or strategies should be adapted to the various contexts. Stakeholders especially recommended carefully assessing the particular needs of rural areas and other underserved areas (e.g. urban slums). Also, the planning for PCPs’ practices should consider the growing urbanisation in Benin and anticipate emerging needs and challenges. Policymakers should consider specific arrangements for the private sector whenever needed. For instance, PCPs from the private sector may need specific financing modalities such as tax reduction, loans, contracts for providing essential health services, etc.

Furthermore, individual PCPs may not be required to embrace all the roles defined in the policy framework (although they should be prepared for them). For example, the various roles can be embraced by a team instead of an individual PCP if there is more than one PCP or other primary care team members better suited for some roles. This is particularly important in Benin, where PCPs work alongside nurse practitioners who are sometimes very experienced and competent in various areas. Similarly, the activities and the organisation of a PCP’s practice can change with the context, as illustrated below:

> *“Now, it also depends on where the doctor is, where they are playing their role as the first line of care. If they are in a remote area where there are no specialists, they become everything, until they can send the patients to where there are specialists, you see? Don’t ask a doctor at the first line in Tchoumi-Tchoumi (a remote area in Benin) who sees a patient with high blood pressure to send them to a cardiologist. They need to give the patient some sort of treatment to help them. When the doctor is sure that the patient can go and see a specialist, they will tell them to go to this or that place so that they can see a specialist…”.* (Local health authority, in-depth interview, Northern Benin, 2022).

## Discussion

### Comparison with international literature and PCPs’ practices in the world

Most of the items included in this policy framework are consistent with the specificities of generalist PCPs’ practices described around the world and key recommendations for strong PHC systems.

However, the Benin PCPs policy framework features several particularities which reflect the country’s context, history, and needs.

#### Objectives assigned to the PCPs in Benin, compared to other settings

The objectives of ensuring quality of care and ensuring a sound referral system and coordination encompass several core competencies expected from the General Practitioner / Family Doctor as outlined by WONCA Europe, including person-centred care, specific problem-solving skills, comprehensive approach, care coordination, etc. [38].

Quality of care and access to services are the two immediate outputs expected from the service delivery within the PHC measurement framework issued by WHO and UNICEF in 2022 [16]. However, though access is usually cited as a key feature of a good PHC system, it is not always clear that the PCPs have a specific responsibility in ensuring this access. The assignment of this objective to the Beninese PCPs aims to draw their attention to the fact that, beyond the clinical aspects, they also have a societal responsibility to contribute to access to care and the community’s well-being. For instance, the PCPs can adopt, and promote among their team, behaviours that will help reduce the costs of care for the patients, such as rational prescription of drugs and lab tests, promoting quality [39].

Regarding the objective to ensure patient and community satisfaction, it strongly reflects a subject currently high on the agenda in Benin [32,40] and the international community [11,41]: improving patient welcome and respectful care. However, this objective could be further discussed in Benin to reflect a more active role for patients and communities. For instance, the WONCA talked of “promoting patient empowerment” [38], and empowering individuals and communities was one of the commitments of the Astana declaration on PHC in 2018 [16].

#### Beninese PCPs’ roles and services organisation versus other countries

Many of the roles proposed for the PCPs in the Benin health system are similar to those described in other African countries [13,42–44] and even beyond [38,45].

The care provider role is obviously found in every description of PCPs’ roles [13,38,44–46]. However, because the number of PCPs in Benin is still limited, stakeholders recommended that PCPs primarily concentrate on the most complex cases or healthcare activities. This is consistent with the views reported elsewhere on family physicians’ role in Africa [11,47], and it entails allowing competent non-physicians to contribute to patient management. This recommendation is also supported by empirical work in Benin, which showed that patients spontaneously chose nurses for simple cases [32]. Furthermore, international literature indicates that task sharing between physicians and nurses increases accessibility [5,39,48] and that experienced nurse practitioners can adequately care for many primary care encounters [49]. However, this recommendation conflicts with the common practice among private PCPs in Benin and the vision of the “médecins généralistes communautaires” [50], which are all geared toward the provision of first-contact care by the PCPs. Moreover, previous experiences with public PCPs in Benin indicated that task-sharing often led many to focus on hospital or administrative work. Therefore, it will be important to find a careful balance between being fully dedicated to first-contact care and only consulting complex cases. Each primary care team can determine the optimal balance based on physician availability, workload, and team members’ expertise while prioritising the community’s access to the best possible quality of care from their first entry into the health system.

The Benin PCPs’ role as guarantors of quality of care is another role widely reported around the world, especially in recent years. Indeed, it is increasingly recognised that PCPs should not only provide individual care but also contribute to the continuous improvement of the quality of the care provided to the community for which they take responsibility [38,43,45,46]. This includes a continuous assessment of the care, followed by corrective actions if necessary [38]. Within this role, the recommendation for the PCPs to train and supervise other primary care team members was most described in the African literature [42–44]. This is not surprising given the recommendations for task sharing with non-physicians and the leadership role assigned to the PCPs.

The role of “interface between populations and other actors” is also described in the literature [38,44,45,51], usually referred to as “coordination” or “synthesis” role. Indeed, the descriptions provided by the stakeholders for this interface role align with WONCA’s recommendations for the synthesis role. This role involves ensuring that patients access the healthcare professionals and technologies needed for their problems appropriately. It also includes protecting the population against unnecessary (usually costly and sometimes harmful) tests, screenings, or treatments, ensuring a good distribution of information, and guiding patients through the health system’s complexity [38].

The leadership role of the PCPs is more and more described for PCPs in Africa [42–44,52] and other settings such as Canada [46]. This leadership role goes beyond the clinical setting. It includes a person-centered leadership able to motivate, collaborate with, and organise all primary care team members around common objectives and values [44,46,52]. As leaders, PCPs are also expected to help the primary care team access the necessary resources to achieve these objectives. This task both supports and requires many other aspects of the policy framework, including community engagement, the PCPs’ role as planners, coordinators and guarantors of quality, etc. Furthermore, while other primary care team members can assume leadership roles [52,53], the performance analysis of PCPs in Benin indicated that granting them leadership responsibilities to PCPs, coupled with the autonomy to exercise them, significantly enhances their performance [32].

The planner and coordinator role seems more controversial. Indeed, with this role, the PCPs in Benin are requested to perform various activities, ranging from organising the work within the health facilities to running health campaigns to conducting purely administrative work. This planning and managerial aspect is explicitly outlined in the job description of public PCPs within a Beninese health region [54]. Similarly, family physicians in various countries [42,43] and “Médecins Généralistes Communautaires” [44] are expected to effectively manage their team and their practices, support their facility with management tasks, and/or organise outreach activities and other public health activities. However, several voices warn against the PCPs’ managerial tasks overwhelming their clinical responsibilities [47]. The Government of Benin also issued an official letter in 2022 requesting the public PCPs to reduce their managerial tasks and dedicate all mornings to patient care.

Therefore, PCPs and healthcare leaders should ensure that administrative tasks stay within reasonable limits. The PCPs’ performance analysis in Benin showed that hiring a professional manager (other than the clinical staff) can greatly enhance the PCPs’ performance and improve resource management [32]. When such a manager is available, the planner role of PCPs can, therefore, be assigned to this manager, or at least the administrative part of this role.

Regarding the organisation of primary care services, stakeholders, including patients and community members, strongly advocate for teamwork within the primary care team in Benin. This aligns with current evidence demonstrating the benefits of teamwork, including better quality of care [16,55], better coordination [38], better access [56], and cost-effectiveness [48]. Teamwork goes beyond simply working together and task-shifting to those considered subordinates [52]. It involves a genuine collaboration where each team member brings an added value. Unfortunately, in many cases, such teamwork is inexistent in Benin [32]. The Benin PCPs policy framework’s recommendation of teamwork will thus improve PCPs’ performance and ensure patients benefit from a diverse range of complementary skills among PCPs and their team members.

Community engagement, another service organisation aspect of the policy framework, is an important attribute described for family medicine and primary care practice in Africa [55,57] and elsewhere [38,45,56]. In Cuba, basic health teams (typically comprising a family doctor, a nurse, and a public health specialist) succeeded in lowering the incidence and prognosis of conditions like hypertension through regular home visits and health education [45]. The number of PCPs in Benin may not (yet) allow all of them to make regular home visits or be permanently present in the community as in Cuba. The community health policy implemented in Benin [58] is thus an excellent opportunity for PCPs as they can work with community health workers to provide the necessary services to their community. However, this should not be an excuse for PCPs to remain in an ivory tower, as is sometimes the case. They need to immerse themselves in their community, understand its social realities, establish regular communication with community leaders, and find mechanisms to remain accessible when needed.

The last aspect of service organisation highlighted by the policy framework is establishing an effective referral system. This referral system would allow a smooth flow of patients across various providers and levels of the health system as needed. A key suggestion is to build referral networks within the primary care level, especially for medical specialists or other cadres (specialised nurses, physiotherapists, social workers, nutritionists, etc.) for whom small facilities cannot provide enough patients to justify their full-time presence. This is already happening in the private sector in Benin, but it is not structured. The polyclinic model in Cuba provides an example of such referrals among primary care facilities [45,59]. These polyclinics are staffed with physicians from various specialities and other health and non-health specialists, offering an average of twenty services, including preventive care, routine curative care, emergency services, rehabilitation, lab tests, etc. [59]. They also serve as an organisational hub for a number of basic health teams, supporting information sharing, coordination of activities, accountability and learning among them [45,51]. The polyclinics were considered key contributors to good results obtained by the Cuban primary health care system [45,51,59]. Also, the rise of polyclinics helped decrease the number of hospitals needed, thus achieving greater efficiency [45,59]. Another example of such primary care hubs is in Brazil with the Family Health Support Centers, which are composed of various specialist physicians, health and social workers, alternative medicine professionals, etc.[56]. However, care must be taken to ensure that opting for polyclinics or similar referral networks does not result in over-medicalising relatively benign health issues and the excessive use of specialised technologies at the primary care level.

Another recommendation concerning the referral system involves thinking beyond the classical primary care centre to district hospital referrals. This entails integrating into the referral networks, after careful analysis, secondary care facilities (for instance, non-profit hospitals or even some private polyclinics) the PCPs already trust and use spontaneously. Indeed, empirical data from a study of 155 PCPs in Benin revealed that 42.6% of them refer emergencies to facilities other than the district hospital, which should formally be the first option. Sometimes, the PCPs’ choice is due to the district hospital’s distance from their facilities. However, in many cases, their choice is driven by a higher level of trust in these other facilities to provide the necessary care for their patients [18]. Whatever referral system is chosen, PCPs and their teams will need direct communication lines, regular contacts for learning and sharing and good coordination with various providers within the referral network. This communication is often lacking in Benin [10], and some good practices can inspire the country. For instance, online platforms can collect and share patient data among relevant providers and provide guidelines and other useful resources to health providers for one-on-one discussions between providers and continuous group discussions [48,60]. Regular learning sessions among providers within the same referral network can also be useful [45,61].

The policy framework proposed in this paper did not provide details on aspects such as opening hours, patient flow management, payment modalities for services by the patients, and other measures aimed at facilitating the primary care team’s work and enhancing the accessibility of PCPs’ practices. It is because such aspects are very specific and should be adapted to the local priorities, the resources available, and the particularities of the community served.

#### Governance of the PCPs’ practices

While the literature often describes the weak governance of the health workforce in sub-Saharan Africa [11,13,62], the descriptions or normative frameworks we found on the PCPs focus on their roles, competencies, values and attributes [38,44,46,63]. Most of the ten governance interventions proposed by the Benin PCPs policy framework are also recommended in various frameworks related to PHC [16] or PHC workforce, including the recent policy document on building PHC teams for UHC in Africa developed by the African Forum for PHC [53]. However, the Beninese framework highlighted elements specific to both the PCPs and the context of Benin.

Developing and adopting adequate regulations and policies is a key intervention, and countries should adopt them for both the public and private PCP [53,64]. This intervention is even more important in Benin, given its huge private sector, where over 90% of PCPs work [18]. Strong regulation can provide PCPs with clear role definitions, adequate resources, and equitable distribution (presently, 90% of PCPs are concentrated in urban areas [18]). Furthermore, adequate regulations can effectively align the PCPs’ work with the national health policy. For instance, posting rules and incentives in Brazil helped improve physicians’ distribution between urban and rural areas [56].

Adequate preparation of the PCPs is the most cited intervention in the literature. This includes undergraduate and postgraduate training adapted to the local context [45,55,56] and providing the PCPs with adequate skills and exposure to general and community practice [42,44,55]. In Benin, this preparation proved crucial as it greatly influences the PCPs’ performance [32] but is often lacking [18].

Providing adequate continuing education [63,64] and establishing a supportive supervision and coaching system [5,32,53] have proven to be pivotal interventions for strengthening the practices of primary care workers, including PCPs. Strangely, there seems to be a prevailing perception that PCPs do not require such interventions, which could explain the current deficiency observed in the continuous education, supervision, and coaching of PCPs in Benin [10,32]. Yet, PCPs, too, require continuous education in both the public and private sectors [64].

Professional organisations can play a significant role in offering this support and coaching for PCPs [5,50], especially because they can reach PCPs from both the private and the public sectors [64]. They can also contribute to regulating their practices and promoting appropriate values. In Benin, most PCPs are affiliated with the National Council of Physicians [18]. However, the functioning of this council needs to be improved. Other professional organisations to which the PCPs are affiliated include the National Association of “Médecins Géneralistes Communautaires”, physicians’ syndicates and private facilities associations for those who own private practices. Unfortunately, only 40% of PCPs are affiliated with any of these organisations (apart from the council) [18]. Enhancing the functioning of these organisations, as suggested by the policy framework, will be essential.

While the literature widely recommends to ensure an appropriate primary care team and adequate equipment, infrastructure, and supplies to support the PCPs’ practices in sub-Saharan Africa [5,45,53], the PCPs performance analysis in Benin [32], along with examples from other countries [45,65] demonstrates that considerable achievements can be made even with limited resources. This is especially possible with robust political support [45], effective team leadership [32,45] and strong partnerships with community members [32,64]. However, this should not prevent from investing to support the work of the primary care teams in Benin.

Financial support is indeed a key recommendation of the policy framework. Studies indicate that practices strongly dependent on self-funding and patient fees often face sustainability challenges [48,66], posing risks to accessibility [32,48], equity [18,48], and even quality of care [32]. These issues are more frequently found in the private sector. Various approaches can help provide financial support to private providers, including tax benefits [64], contracting private providers for some aspects of service delivery [48,64], or financial support for initial practice start-ups [50]. Moreover, non-financial support and advice can be provided to PCPs and their teams, aiming to reduce healthcare production costs without compromising quality. This can be, for instance, social franchising for sharing administrative costs and getting more credibility [48], giving the PCPs access to a professional facility manager [32], supporting them in establishing their business plan [50], promoting the use of generic drugs [45,48], using diagnostic support software [39,48,64], etc. Benin is building its national health insurance scheme and considering contracting with private providers. This presents an opportunity for private PCPs as these contracts could bolster their financial sustainability. In return, it can assist the government in regulating their practices effectively and better aligning them with national health objectives.

Health workers’ accountability is increasingly mentioned in papers on health workforce development and quality of care. Several frameworks, for instance, the CanMEDS 2015 physician competency framework [46] or the Statement of Consensus on Family Medicine in Africa [63], clearly establish the social accountability of the PCPs. However, few documents explicitly state how to achieve this accountability. Usually, only some aspects of accountability are explicated, mostly the clear definition of roles and responsibilities [53] and a clear definition of the population of responsibility [53,57,64]. Similarly, in Benin, only some aspects of accountability are currently implemented. There is a job description for the PCPs working in the public sector in Benin, and the population for whom they are responsible is relatively well-defined, determined by the geographic area assigned to their health facility [54]. However, job descriptions are not consistently present in the private sector, and there is almost no formal definition of the population for which private PCPs are responsible.

Consequently, they barely feel responsible for monitoring the health status of a given population or addressing the needs of a specific community. Additionally, when public and private facilities share the same geographic area (especially in urban settings), attributing responsibilities can be challenging when a problem arises. Private PCPs in Benin seldom participate in monitoring local health system indicators. As for regular assessments of PCP practices and accountability platforms to report to the populations, they are insufficiently implemented in both the public and private sectors in Benin. The Benin PCPs policy framework provides recommendations that can help holistically address accountability issues.

The benefits of an effective health information system to support the PCPs’ performance are largely shown. For instance, in Cuba, family doctors and their teams are required to gather and analyse data on their assigned community and present the results at local meetings with community groups [45,51]. In Belgium, the Intego database [67] relies on electronic health records to routinely compile a large amount of data on the PCPs’ practices. This data helps to understand and detect changes in the epidemiological profile of the population. The data also helps analyse the PCPs’ practices (drug prescription, for example) [67]. In Benin, the health information system mainly collects data (usually aggregated for each facility) on key health indicators, which is helpful in following the progress on these indicators at the national and sub-national levels. However, the health information system should provide detailed information on individual patients and care processes for a regular analysis of the care practices and changes in the population’s needs. Moreover, the patient files in Benin are still largely paper-based [18], with risks of data loss or errors during reporting. It also requires a lot of extraction work and resources if one wants to assess the care process for quality improvement or research purposes. Digitalising the health information system at the facility level, with interconnection to national databases, can help gather detailed health information and support data analysis [48,67]. It is feasible in Benin despite the still limited electricity and computer access.

Indeed, offline solutions exist and can be adapted for use on smartphones, and the mobile internet penetration in Benin [68] was 68,92% in 2022 (likely higher among health workers).

The Benin PCPs policy framework focuses on enhancing routine information systems rather than placing a significant emphasis on research, unlike other countries [42,46]. This lesser emphasis on research does not imply a lack of recognition that PCPs should engage in relevant research for their practice and community. Instead, it reflects a deliberate focus on establishing first a conducive environment with accessible and reliable data.

#### Professional identity and contextual aspects

The Benin PCPs policy framework outlines seven fundamental values for PCPs and other primary care workers: respect for people, empathy, altruism, humility, professionalism, community orientation, and integrity. These values align with recognised core values for PCPs in Africa [13] and other regions [38], and many training programmes [38,42,46,69] address them. However, PCPS need to work within environments that nurture these values. Indeed, despite their best intentions, some PCPs find themselves in situations that conflict with the values they are supposed to embrace [32,70]. Hence, there is a need to establish a strong professional identity for PCPs. Building this professional identity requires an official status for PCPs. The latter can also help to make primary care attractive to young physicians, develop relevant training curricula, and better organise these practices [38,42].

Adapting health interventions and policies to the local context is a well-known principle in global health [36,38,55]. For instance, because of its limited resources and access to medicines, Cuba had to develop its own training for PCPs, its own protocols of care, and its own care organisation [45,59]. However, as tempting as it may be to copy the Cuban model because of its successes, it would be a mistake for Benin, given the two countries’ very different political, social, historical, and cultural contexts. Similarly, given that Benin’s resources are far from those of Western countries, copying and pasting models in these contexts is not an option either. Therefore, while integrating lessons from other settings, the Benin policy framework emphasises core elements related to PCP practices in Benin, as deemed relevant by multiple stakeholders and supported by context-related evidence. We also provided concrete examples of how these elements can be applied. However, as largely shown in previous paragraphs, several contextual specificities will require adaptation and sometimes choices among the various options proposed by the policy framework. This, in turn, requires sound leadership, a sufficient level of autonomy for PCPs, their team and local health authorities and a rigorous learning process [71]. Other context-related issues must be discussed further to see how the PCPs can adapt or cope with them. This is, for instance, the case of the wide use of alternative care providers in Benin [72] or the rapid population growth and urbanisation [73], resulting in the persistence or emergence of complex health problems. Finally, the policy framework should be regularly evaluated and updated, as is the case in many other contexts [38,45,51,56].

### Strengths and limitations

The policy framework presented in this paper was developed through a meticulous cocreation process, using empirical data and analysis from multiple stakeholders. Moreover, we carefully compared the elements of the policy framework with existing international experiences, allowing for critical analysis to evaluate their scientific basis and feasibility.

In terms of limitations, the most obvious is that this policy framework is highly contextualised, needing adaptations for application to other countries. However, given the scientific basis underpinning it and that other African countries share similarities with Benin, the framework can serve as a valuable starting point for countries looking to guide PCPs’ practices. Furthermore, the West African regional context is currently undergoing profound changes, including the rise of terrorism, repeated *coups d’Etat* and an increasingly fragile socio-economic situation. The policy framework, as it stands, considers empirical data up to 2022 and the context up to 2023. It is difficult to predict how current geopolitical and socio-economic changes will impact the sustainability of its recommendations. Thus, we emphasise the importance of implementing the framework alongside continuous learning processes to ensure adaptability as circumstances evolve.

## Conclusion

The cocreation process led to an evidence-informed and consensus-based policy framework for guiding PCPs’ practices in Benin. This framework provides a roadmap to ensure a better contribution of PCPs to the improvement of Benin’s PHC system compared to the current situation. The various interventions and recommendations presented in this framework must be tested, and a learning process is needed for continuous improvement. While the focus of the policy framework is on the case of Benin, the findings of the empirical work supporting it, the interventions recommended and many other aspects are of relevance to other countries facing similar contexts (low to lower-middle income, high mortality and morbidity, unregulated and poorly supported medical practices, need to improve the quality of care with limited resources, etc.), especially in francophone West Africa.

## Data Availability

All relevant data are within the manuscript and its Supporting Information files.

## Acknowledgements

Our gratitude goes first to all participants in the various studies and activities that supported the cocreation process. We also thank the participants in the cocreation workshop for their useful reflections and the precious time they allocated to the development of the policy framework. We want to acknowledge the assistance provided by Harziki BOUKARI, Astrid FLENON, Cybèle HOUNMENOU, and Armelle VIGAN GUEZODJE, who played pivotal roles in facilitating the workshop. Finally, we are grateful to Vanessa SEKPON for her invaluable support in facilitating and synthesising the workshop’s data.

## Supporting information captions

S1 file: report of the cocreation workshop

S2 file: Combination empirical and workshop data

## References

1. World Health Organization. The Global Health Observatory [cited 27 December 2023]. Available from: https://www.who.int/data/gho/data/indicators/.

2. Macinko J, Starfield B, Erinosho T. The impact of primary healthcare on population health in low- and middle-income countries. J Ambul Care Manage. 2009;32: 150–71. doi: 10.1097/JAC.0b013e3181994221.

3. World Health Organization. Primary health care [cited 22 September 2023]. Available from: https://www.who.int/health-topics/primary-health-care#tab=tab_1.

4. Kringos DS, Boerma WGW, Hutchinson A, van der Zee J, Groenewegen PP. The Breadth of primary care. BMC Health Serv Res. 2010;10: 65. doi: 10.1186/1472-6963-10-65.

5. World Health Organization. Global strategy on human resources for health: Workforce 2030. Geneva: World Health Organization; 2016. Available from: https://iris.who.int/bitstream/handle/10665/250368/9789241511131-eng.pdf?sequence=1

6. Eyal N, Cancedda C, Kyamanywa P, Hurst SA. Non-physician clinicians in sub-Saharan Africa and the evolving role of physicians. Int J Health Policy Manag. 2015;5(3):149–53. doi: 10.15171/ijhpm.2015.215.

7. Willcox ML, Peersman W, Daou P, Diakité C, Bajunirwe F, Mubangizi V, et al. Human resources for primary health care in sub-Saharan Africa: progress or stagnation? Hum Resour Health. 2015;13(76):1–11. doi: 10.1186/s12960-015-0073-8

8. Direction de la programmation et de la prospective. Annuaire Statistiques Sanitaires du Bénin 2017. Cotonou: Ministère de la santé du Bénin; 2018.

9. Direction de la programmation et de la prospective. Annuaire des statistiques sanitaires 2010 du Bénin. Cotonou: Ministère de la Santé du Bénin; 2011.

10. Bello K, De Lepeleire J, Agossou C, Apers L, Zannou DM, Criel B. Lessons learnt from the experiences of primary care physicians facing COVID-19 in Benin: A mixed-methods study. Frontiers in Health Services. 2022;2: 11. doi: 10.3389/frhs.2022.843058.

11. Mash R, Howe A, Olayemi O, Makwero M, Ray S, Zerihun M, et al. Reflections on family medicine and primary healthcare in sub-Saharan Africa. BMJ Glob Health. 2018;3: e000662. doi: 10.1136/bmjgh-2017-000662.

12. Von Pressentin KB, Mash RJ, Baldwin-Ragaven L, Botha RPG, Govender I, Steinberg WJ. The bird’s-eye perspective: how do district health managers experience the impact of family physicians within the South African district health system? A qualitative study. South African Fam Pract. 2018;60(1):13–20. doi: 10.1080/20786190.2017.1348047.

13. Bello K, De Lepeleire J, Kabinda M. J, Bosongo S, Dossou J-P, Waweru E, et al. The expanding movement of primary care physicians operating at the first line of healthcare delivery systems in sub-Saharan Africa: A scoping review. PLoS One. 2021;16: e0258955. doi: 10.1371/journal.pone.0258955.

14. Jan T, Aillet S. De Tananarive au Caire : un aperçu du métier de médecin généraliste libéral dans le contexte des systèmes de santé locaux [Thèse de Médecine]. Bordeaux: Université Bordeaux 2 – Victor Segalen UFR des sciences médicales; 2011. Available from : https://bdsp-ehesp.inist.fr/vibad/index.php?action=getRecordDetail&idt=433735.

15. Bosongo SI, Mukalenge FC, Tambwe AM, Criel B. Les médecins prestataires à la première ligne des soins dans la ville de Kisangani en République Démocratique du Congo : vers une typologie. Afr J Prim Health Care Fam Med. 2021;13: 8. doi: 10.4102/phcfm.v13i1.2617.

16. Word Health Organization, the United Nations Children’s Fund. Primary health care measurement framework and indicators: monitoring health systems through a primary health care lens. Geneva: World Health Organization; 2022. Available from: https://iris.who.int/bitstream/handle/10665/352205/9789240044210-eng.pdf?sequence=1.

17. Patnaik P. A policy framework towards the use of artificial intelligence by public institutions: Reference to FATE analysis. In: Saura JR, Debasa F, editors. Handbook of Research on Artificial Intelligence in Government Practices and Processes. Hershey IGI Global; 2022. doi: 10.4018/978-1-7998-9609-8.ch002.

18. Criel B, Bello K, Tawaytibhongs O, Waweru E. The growing presence of medical doctors at first line healthcare level in Benin, Thailand and Uganda: a multifaceted phenomenon. Berlin: The European Conference of Family Doctors; 2020.

19. Hansen PM, Synowiec C, Blanchet NJ. Co-production between researchers and policymakers is critical for achieving health systems change. 2021 Feb 15 [cited 28 June 2023]. In: thebmjopinion [Internet]. BMJ Journals: 2021. Available from https://blogs.bmj.com/bmj/2021/02/15/co-production-between-researchers-and-policymakers-is-critical-for-achieving-health-systems-change/.

20. Adams E, Bello K, Blanchet N, Dossou J-P, Issa A, Miller L, et al. How Togo recharted its path to universal health coverage amid COVID-19 [cited 26 September 2023]. Washington: 2021. Available from: https://www.acceleratehss.org/2021/01/22/how-togo-recharted-its-path-to-universal-health-coverage-amid-covid-19.

21. Redman S, Greenhalgh T, Adedokun L, Staniszewska S, Denegri S. Co-production of knowledge: the future. BMJ. 2021;372 : n434. doi: 10.1136/bmj.n434.

22. Rajan D, Adam T, El Husseiny D, Porignon D, Ghaffar A, Schmets G. Policy dialogue : What it is and how it can contribute to evidence-informed decision-making. Geneva: World Health Organization; 2015. Available from: https://extranet.who.int/uhcpartnership/sites/default/files/reports/2015-Briefing-Note.pdf.

23. Partridge ACR, Mansilla C, Randhawa H, Lavis JN, El-Jardali F, Sewankambo NK. Lessons learned from descriptions and evaluations of knowledge translation platforms supporting evidence-informed policy-making in low- and middle-income countries: a systematic review. Health Res Policy Syst. 2020;18: 127.doi: 10.1186/s12961-020-00626-5.

24. Shroff Z, Aulakh B, Gilson L, Agyepong IA, El-Jardali F, Ghaffar A. Incorporating research evidence into decision-making processes: researcher and decision-maker perceptions from five low- and middle-income countries. Health Res Policy Syst. 2015;13: 70. doi: 10.1186/s12961-015-0059-y.

25. Robert E, Rajan D, Koch K, Weaver AM, Porignon D, Ridde V. Policy dialogue as a collaborative tool for multistakeholder health governance: a scoping study. BMJ Glob Health. 2020;4: e002161. doi: 10.1136/bmjgh-2019-002161.

26. Marten R, El-Jardali F, Hafeez A, Hanefeld J, Leung GM, Ghaffar A. Co-producing the covid-19 response in Germany, Hong Kong, Lebanon, and Pakistan. BMJ. 2021;372: n243. doi: 10.1136/bmj.n243 .

27. Loffreda G, Bello K, Kiendrébéogo JA, Selenou I, Ag Hamed MA, Dossou JP, et al. Political economy analysis of universal health coverage and health financing reforms in low-and middle- income countries: the role of stakeholder engagement in the research process. Health Res Policy Sys. 2021;19: 143.doi: 10.1186/s12961-021-00788-w.

28. Ridde V, Dagenais C. What we have learnt (so far) about deliberative dialogue for evidence-based policymaking in West Africa. BMJ Glob Health. 2017;2: e000432. doi: 10.1136/bmjgh-2017-000432.

29. Dovlo D, Nabyonga-orem J, Estrelli Y, Mwisongo A. Policy dialogues – the “bolts and joints” of policy-making : experiences from Cabo Verde , Chad and Mali. BMC Health Serv Res. 2016;16: 216. doi: 10.1186/s12913-016-1455-x.

30. Meessen B, Akhnif ELH, Kiendrébéogo JA, Belghiti Alaoui A, Bello K, Bhattacharyya S, et al. Learning for Universal Health Coverage. BMJ Glob Health. 2019;4(6): e002059.. doi: 10.1136/bmjgh-2019-002059.

31. El-Jardali F, Lavis J, Moat K, Pantoja T, Ataya N. Capturing lessons learned from evidence-to- policy initiatives through structured reflection. Health Res Policy Sys. 2014;12: 1–15. doi: 10.1186/1478-4505-12-2.

32. Bello K, De Lepeleire J, Zannou DM, Criel B. Factors supporting the primary care physicians’ performance in Benin: a multiple case study. Cotonou: 2023.

33. Delbecq AL, Van de Ven AH. A Group process model for problem identification and program planning. J Appl Behav Sci. 1971;7: 466–92. 10.1177/002188637100700404.

34. Donabedian A. Evaluating the quality of medical care. Milbank Q. 2005;83: 691–729. doi: 10.1111/j.1468-0009.2005.00397.x

35. World Health Organization. Everybody’s business: Strengthening health systems to improve health outcomes: WHO’s framework for action. Geneva: World Health Organization; 2007. Available from: https://iris.who.int/bitstream/handle/10665/43918/9789241596077_eng.pdf?sequence=1.

36. Van Olmen J, Criel B, Bhojani U, Marchal B, Van Belle S; Chenge MF, et al. The health system dynamics framework: the introduction of an analytical model for health system analysis and its application to two case-studies. Health Cult Soc. 2012;2(1):1–21. doi: 10.5195/HCS.2012.71.

37. Caza BB, Creary SJ. The construction of professional identity. In: Wilkinson A, Hislop D, Coupland C, editors. Perspectives on contemporary professional work: Challenges and experiences. Cheltenham :Edward Elgar Publishing; 2016. p. 259–85. https://hdl.handle.net/1813/71987.

38. Wonca Europe. The European definition of general practice / family medicine- 2023 Edition. Ljubljana: Wonca Europe; 2023. Available from: https://www.woncaeurope.org/file/41f61fb9-47d5-4721-884e-603f4afa6588/WONCA_European_Definitions_2_v7.pdf.

39. Mills A, Palmer N, Gilson L, McIntyrec D, Schneider H, Sinanovic E, et al. The performance of different models of primary care provision in Southern Africa. Soc Sci Med. 2004 Sep;59(5):931–43. DOI: 10.1016/j.socscimed.2003.12.015.

40. Maison de la Société Civile du Bénin. Pétition pour l’amélioration des services de santé au Bénin : des soins de qualité et l’accès aux médicaments essentiels. 2023 June 15 [Cited 18 November 2023]. Cotonou: 2023. Available from: https://www.mdscbenin.org/sante/.

41. Global Respectful Maternity Care Council. Respectful maternity care: The universal rights of women & newborns. Washington: White Ribbon Alliance; 2019. Available from: https://whiteribbonalliance.org/wp-content/uploads/2022/05/WRA_RMC_Charter_FINAL.pdf

42. Mash R. Reflections on the development of family medicine in the Western Cape: a 15-year review. South African Fam Pract. 2011;53(6):557–62. doi: 10.1080/20786204.2011.10874152.

43. Besigye IK, Onyango J, Ndoboli F, Hunt V, Haq C, Namatovu J. Roles and challenges of family physicians in Uganda: a qualitative study. Afr J Prim Health Care Fam Med. 2019;11(1):1–9. doi: 10.4102/PHCFM.V11I1.2009.

44. Desplats D, Razakarison C. Le guide du Médecin Généraliste Communautaire. Marseille: Santé Sud; 2011. Available from : https://mediatheque.agencemicroprojets.org/wp-content/uploads/Guide-medecin-generaliste-communautaire.pdf.

45. Pineo R. Cuban Public Healthcare: A model of success for developing nations. J Dev Soc. 2019;35: 16–61. 10.1177/0169796X19826731.

46. Frank JR, Snell L, Sherbino J, editors. CanMEDS 2015 Physician Competency Framework. Ottawa: Royal College of Physicians and Surgeons of Canada; 2015. Available from: https://canmeds.royalcollege.ca/uploads/en/framework/CanMEDS%202015%20Framework_EN_Reduced.pdf.

47. Moosa S, Downing R, Essuman A, Pentz S, Reid S, Mash R. African leaders’ views on critical human resource issues for the implementation of family medicine in Africa. Hum Resour Health. 2014 Jan;12(2):1–9. doi: 10.1186/1478-4491-12-2.

48. Girdwood S, Govender K, Long L, Miot J, Meyer-Rath G. Primary healthcare delivery models for uninsured low-income earners during the transition to National Health Insurance: perspectives of private South African providers. S Afr Med J. 2019 Sep;109(10):771–83. doi: 10.7196/SAMJ.2019.v109i10.13930.

49. Christoffels R, Mash R. How well do public sector primary care providers function as medical generalists in Cape Town: a descriptive survey. BMC Fam Pract. 2018 Jul;19(122): 1–9. doi: 10.1186/s12875-018-0802-x.

50. Desplats D, Koné Y, Razakarison C. Pour une médecine générale communautaire en première ligne. Med Trop. 2004;64(6):539–44. French. DOI: 10.1055/s-0029-1237558.

51. Reed G. Cuba’s primary health care revolution: 30 years on. Bull World Health Organ. 2008;86(5): 327–9. doi: 10.2471/blt.08.030508.

52. Moosa S. Family doctor leadership in African primary health care. Afr J Prim Health Care Fam Med 2021;13: 1–2. doi: 10.4102/phcfm.v13i1.3198.

53. The African Forum for primary health care. Building effective primary health care teams for UHC in Africa. Johannesburg: 2023. Available from : https://afrophc.org/category/policy-framework/

54. Direction Départementale de la Santé du Borgou. Fiches de description de poste des acteurs de la périphérie : Poste : médecin responsable du centre de santé. Parakou : DDS Borgou; 2022.

55. Moosa S. WONCA Africa: Introduction to family doctors [cited 1 March 2024]. Johannesburg: 2020. Available from: https://profmoosa.com/wp-content/uploads/2020/02/wonca-africa-flyer.pdf.

56. OECD. Health care needs and organisation of primary health care in Brazil. In: Primary Health Care in Brazil. Paris: OECD Reviews of Health Systems, OECD Publishing. 10.1787/120e170e-en.

57. Mash R, Ray S, Essuman A, Burgueño E. Community-orientated primary care: a scoping review of different models, and their effectiveness and feasibility in sub-Saharan Africa. BMJ Glob Health. 2019;4: e001489. doi: 10.1136/bmjgh-2019-001489.

58. Ministère de la santé du Bénin. Politique Nationale de la Santé Communautaire. Cotonou : Ministère de la santé du Bénin; 2015. Available from : https://www.medbox.org/document/politique-nationale-de-la-sante-communautaire#GO.

59. Lamrani S. The Health System in Cuba: Origin, Doctrine and Results. Études caribéennes. 2021;6. doi: 10.4000/etudescaribeennes.24110.

60. Delvaux N, Piessens V, De Burghgraeve T, Mamouris P, Vaes B, Vander Stichele R, et al. Clinical decision support improves the appropriateness of laboratory test ordering in primary care without increasing diagnostic error: the ELMO cluster randomised trial. Implementation Sci. 2020;15(100). doi: 10.1186/s13012-020-01059-y.

61. Fernandes Q, Augusto O, Uetela DM, Sherr K. Mozambique: Addressing neonatal mortality through a peer-to-peer learning intervention at district level. In: Sheikh K, Abimbola S, editors. Learning health systems: pathways to progress. Flagship report of the Alliance for Health Policy and Systems Research. Geneva: World Health Organization; 2021. Available from: https://iris.who.int/bitstream/handle/10665/344891/9789240032217-eng.pdf?sequence=1.

62. Rice J, Hunter M, Kimathi G, Mukami D, Hetta R, Ntwiga I. Health workforce governance: Quality an essential ingredient for enhanced health sector workforces in Africa. Nairobi: AMREF Africa; 2023. Available from: https://repository.amref.ac.ke/bitstream/handle/20.500.14173/843/Quality%20An%20Essential%20Ingredient%20for%20Enhanced%20Health%20Sector%20Workforces%20in%20Africa.pdf?sequence=1&isAllowed=y

63. Mash R, Reid S. Statement of consensus on Family Medicine in Africa. Afr J Prim Health Care Fam Med 2010;2: 4–7. doi: 10.4102/phcfm.v2i1.151.

64. World Health Organization, United Nations Children’s Fund. Operational framework for primary health care: Transforming vision into action. Geneva: World Health Organization and the United Nations Children’s Fund; 2020. Available from: https://iris.who.int/bitstream/handle/10665/337641/9789240017832-eng.pdf?sequence=1.

65. Nimpagaritse M, Korachais C, Meessen B. Effects in spite of tough constraints - A theory of change based investigation of contextual and implementation factors affecting the results of a performance based financing scheme extended to malnutrition in Burundi. PLoS One. 2020;15(1): e0226376. doi: 10.1371/journal.pone.0226376.

66. Hellowell M, Myburgh A, Sjoblom M, Gurazada S, Clarke D. Covid-19 and the collapse of the private health sector: a threat to countries’ response efforts and the future of health systems strengthening? Geneva: Health Systems Governance Collaborative; 2020. Available from: https://hsgovcollab.org/en/blog/covid-19-and-collapse-private-health-sector-threat-countries-response-efforts-and-future.

67. Truyers C, Goderis G, Dewitte H, Akker M Vanden, Buntinx F. The Intego database: Background, methods and basic results of a Flemish general practice-based continuous morbidity registration project. BMC Med Inform Decis Mak. 2014;14(48). doi: 10.1186/1472-6947-14-48.

68. Autorité de Régulation des Communications Électroniques et de la Poste (ARCEP). Observatoire de l’Internet au Bénin : Tableau de bord au 31 Mars 2022. Cotonou: Arcep Bénin; 2022. Available from : https://arcep.bj/wp-content/uploads/2022/06/Tableau-de-bord-Internet-au-31-Mars-2022.pdf.

69. Van Dormael M. L’introduction des sciences sociales dans une expérience de formation professionnelle des médecins de campagne au Mali. Colloque International Amades : Proceedings of the international colloquium of Anthropology and Medicine; 2007 Oct 25-27; Marseille, France. Available from : http://www.strengtheninghealthsystems.be/doc/3/ref%203.6%20=%20ref%202.14%20introduction%20des%20sciences%20sociales%20-%20Mali.pdf.

70. Van der Voort CTM, Van Kasteren G, Chege P, Dinant GJ. What challenges hamper Kenyan family physicians in pursuing their family medicine mandate? A qualitative study among family physicians and their colleagues. BMC Fam Pract. 2012 Apr;13(32):1–15 DOI: 10.1186/1471-2296-13-32.

71. Sheikh K, Abimbola S, editors. Learning health systems: pathways to progress. Flagship report of the Alliance for Health Policy and Systems Research. Geneva: World Health Organization; 2021. Available from: https://iris.who.int/bitstream/handle/10665/344891/9789240032217-eng.pdf?sequence=1.

72. Gryseels C, Dossou JP, Vigan A, Boyi Hounsou C, Kanhonou L, Benova L, et al. Where and why do we lose women from the continuum of care in maternal health? A mixed-methods study in Southern Benin. Tropical Medicine and International Health 2022;27: 236–43. doi: 10.1111/tmi.13729.

73. The World Bank. DataBank: World Development Indicators [cited 09 August 2024]. Available from: https://databank.worldbank.org/reports.aspx?dsid=2&series=SP.URB.TOTL.IN.ZS.

